# A citizen science initiative for open data and visualization of COVID-19 outbreak in Kerala, India

**DOI:** 10.1101/2020.05.13.20092510

**Authors:** Jijo Pulickiyil Ulahannan, Nikhil Narayanan, Nishad Thalhath, Prem Prabhakaran, Sreekanth Chaliyeduth, Sooraj P Suresh, Musfir Mohammed, E Rajeevan, Sindhu Joseph, Akhil Balakrishnan, Jeevan Uthaman, Manoj Karingamadathil, Sunil Thonikkuzhiyil Thomas, Unnikrishnan Sureshkumar, Shabeesh Balan, Neetha Nanoth Vellichirammal

## Abstract

**Objective:** India reported its first COVID-19 case in the state of Kerala and an outbreak initiated subsequently. The Department of Health Services, Government of Kerala, initially released daily updates through daily textual bulletins for public awareness to control the spread of the disease. However, this unstructured data limits upstream applications, such as visualization, and analysis, thus demanding refinement to generate open and reusable datasets.

**Materials and Methods:** Through a citizen science initiative, we leveraged publicly available and crowd-verified data on COVID-19 outbreak in Kerala from the government bulletins and media outlets to generate reusable datasets. This was further visualized as a dashboard through a frontend web application and a JSON repository, which serves as an API for the frontend.

**Results:** From the sourced data, we provided real-time analysis, and daily updates of COVID-19 cases in Kerala, through a user-friendly bilingual dashboard (https://covid19kerala.info/) for non-specialists. To ensure longevity and reusability, the dataset was deposited in an open-access public repository for future analysis. Finally, we provide outbreak trends and demographic characteristics of the individuals affected with COVID-19 in Kerala during the first 138 days of the outbreak.

**Discussion:** We anticipate that our dataset can form the basis for future studies, supplemented with clinical and epidemiological data from the individuals affected with COVID-19 in Kerala.

**Conclusion:** We reported a citizen science initiative on the COVID-19 outbreak in Kerala to collect and deposit data in a structured format, which was utilized for visualizing the outbreak trend and describing demographic characteristics of affected individuals.

## BACKGROUND AND SIGNIFICANCE

In December 2019, an outbreak of cases presenting with pneumonia of unknown etiology was reported in Wuhan, China. The outbreak, caused by a novel severe acute respiratory syndrome Coronavirus-2 (SARS-CoV-2), later evolved as a pandemic (coronavirus disease 2019; COVID-19), claiming thousands of lives globally. [1-4] Initial studies revealed the clinical and prognostic features of COVID-19 along with its transmission dynamics and stressed the need for implementing public health measures for containment of infection and transmission among the population at high-risk. [2 5-9] In response to this, several countries have implemented measures including travel restrictions and physical distancing by community-wide quarantine. [2 6 10] These extensive measures were imposed, taking into consideration the lack of adequate testing kits for detection, a vaccine, or proven antivirals for preventing or treating this disease along with reports of considerable strain on the health system leading to unprecedented loss of human life.

India—the second most populated country in the world—reported its first case in the state of Kerala on January 30, 2020, among individuals with travel history from Wuhan, the epicenter of the COVID-19 outbreak. [11] With the subsequent reports of an outbreak in the Middle East and Europe, Kerala has been on high-alert for a potential outbreak, as an estimated 10% of the population work abroad and being an international tourist destination. [12 13] The state has a high population density, with a large proportion of the population falling in the adult and older age group. [14] This population also shows a high incidence of COVID-19-associated comorbidities such as hypertension, diabetes, and cardiovascular disease. [9 15-17] As evidenced by reports of other countries, these factors pose a significant threat for an outbreak and would exert a tremendous burden on the public healthcare system. [18-20]

Severe public health measures were implemented in the state of Kerala and across India to prevent an outbreak. International flights were banned by March 22, 2020, and a nation-wide lockdown was initiated on March 25, 2020. [21] However, before these measures were implemented, several cases (including travelers from Europe and the Middle East), along with a few reports of secondary transmission, were reported in Kerala. Since the first case was reported, the Department of Health Services (DHS), Government of Kerala, initiated diagnostic testing, isolation, contact tracing, and social distancing through quarantine, and the details of cases were released for the public through daily textual bulletins.

For pandemics such as COVID-19, public awareness via dissemination of reliable information in real-time plays a significant role in controlling the spread of the disease. Besides, real-time monitoring for identifying the magnitude of spread helps in hotspot identification, potential intervention measures, resource allocation, and crisis management. [22] The lack of such a real-time data visualization dashboard for the public with granular information specific to Kerala in the local language (Malayalam), during the initial days of the outbreak, was the motivation for this work.

To achieve this, the collection of relevant information on infection and refining the dataset in a structured manner for upstream purposes such as visualization and/or epidemiological analysis is essential. Open or crowd-sourced data has immense potential during the early stage of an outbreak, considering the limitation of obtaining detailed clinical and epidemiological data in real-time during an outbreak. [23-25] Furthermore, the structured datasets, when deposited in open repositories and archived, can ensure longevity for future analytical efforts and policymaking. The unavailability of such structured, reusable, and crowd-verified datasets on natural disasters in Kerala, documented in the public domain, also motivated us to generate a resource for the COVID-19 outbreak. This initiative was volunteered by the Collective for Open Data Distribution-Keralam (CODD-K), a group of technologists, academicians, students, and the public advocating for open data. This collective, in a primitive form, was initiated during the devastating 2018 Kerala floods, which brought together the experts and general public through social media platforms to coordinate rescue missions through citizen-led open/crowd-sourcing strategies.

Here, we report a citizen science initiative to leverage publicly available data on COVID-19 cases in Kerala from the daily bulletins released by the DHS, Government of Kerala, and various news outlets. The multi-sourced data was refined to make a structured live dataset to provide real-time analysis and daily updates of COVID-19 cases in Kerala through a bilingual (English and Malayalam) user-friendly dashboard (https://covid19kerala.info/). We aimed to disseminate the data of the outbreak trend, hotspots maps, and daily statistics in a comprehensible manner for non-specialists with bilingual (Malayalam and English) interpretation. Next, we aimed for the longevity and reusability of the datasets by depositing it in a public repository, aligning with open data principles for future analytical efforts. [26] Finally, to show the scope of the sourced data, we provide a snapshot of outbreak trends and demographic characteristics of the individuals affected with COVID-19 in Kerala during the first 138 days of the outbreak.

## METHODS

### The citizen-led collective for data sourcing and curation

The CODD-K constituting, members from different domains, who shared the interest for sourcing data, building the dataset, visualizing, distributing, and interpreting the data on infection outbreak volunteered this effort (https://team.covid19kerala.info/). This initiative was in agreement with definitions proposed by different citizen-science initiatives.[27 28] The CODD-K invited participation in this initiative from the public through social media. The domain experts in the collective defined the data of interest to be collected, established the informatics workflow, and the web application for data visualization. The volunteers contributed by sourcing data from various media outlets for enriching the data. Dedicated social media (dedicated Telegram channels and WhatsApp groups) channels were used for data collection, which was verified independently and curated by data validation team members.

### Definition and Scope of Datasets

The collective defined the data of interest as minimal structured metadata of the COVID-19 infections in Kerala, covering the possible facets of its spatial and temporal nature, excluding the clinical records (Supplementary Methods). The resulting datasets should maintain homogeneity and consistency, assuring the privacy and anonymity of the individuals. The notion of this data definition is to make the resulting datasets reusable and interoperable with similar or related datasets. A set of controlled vocabularies were formed as a core of this knowledge organization system to reduce anomalies, prevent typographical errors, and duplicate entries. Together with the controlled vocabularies, identifiers of individual entries in each dataset make the datasets interlinked. An essential set of authority control is used in populating spatial data to make it accurate in the naming and hierarchy. A substantial set of secondary datasets were also produced and maintained along with the primary datasets, including derived and combined information from the primary datasets and external resources.

### Data Collection

We primarily sourced publicly available de-identified data, released daily as textual bulletins (from January 31, 2020) by the DHS, Government of Kerala, India (https://dhs.kerala.gov.in), of the individuals diagnostically confirmed positive for SARS-CoV-2 by reverse transcription-polymerase chain reaction (RT-PCR) at the government-approved test centers. We also collected and curated reports from print and visual media for supplementing the data (Supplementary methods). The quality of the data in terms of veracity and selection bias has been ensured as described (Supplementary Methods). Utmost care was taken to remove any identifiable information to ensure the privacy of the subjects. Entries were verified independently by CODD-K data validation team members and rectified for inconsistencies (Figure 1). Since the data collected were publicly available, no individual consent and ethical approval were required for the study. To demonstrate the utility of the collected dataset, we provided the status of the first 138 days (between January 30, 2020, and June 15, 2020) of the COVID-19 outbreak in Kerala, and also described demographic characteristics of the individuals affected with COVID-19. We ensured that the sourced dataset complied with the Open Definition 2.1 laid down by Open Knowledge Foundation. [26]

**Figure 1:**
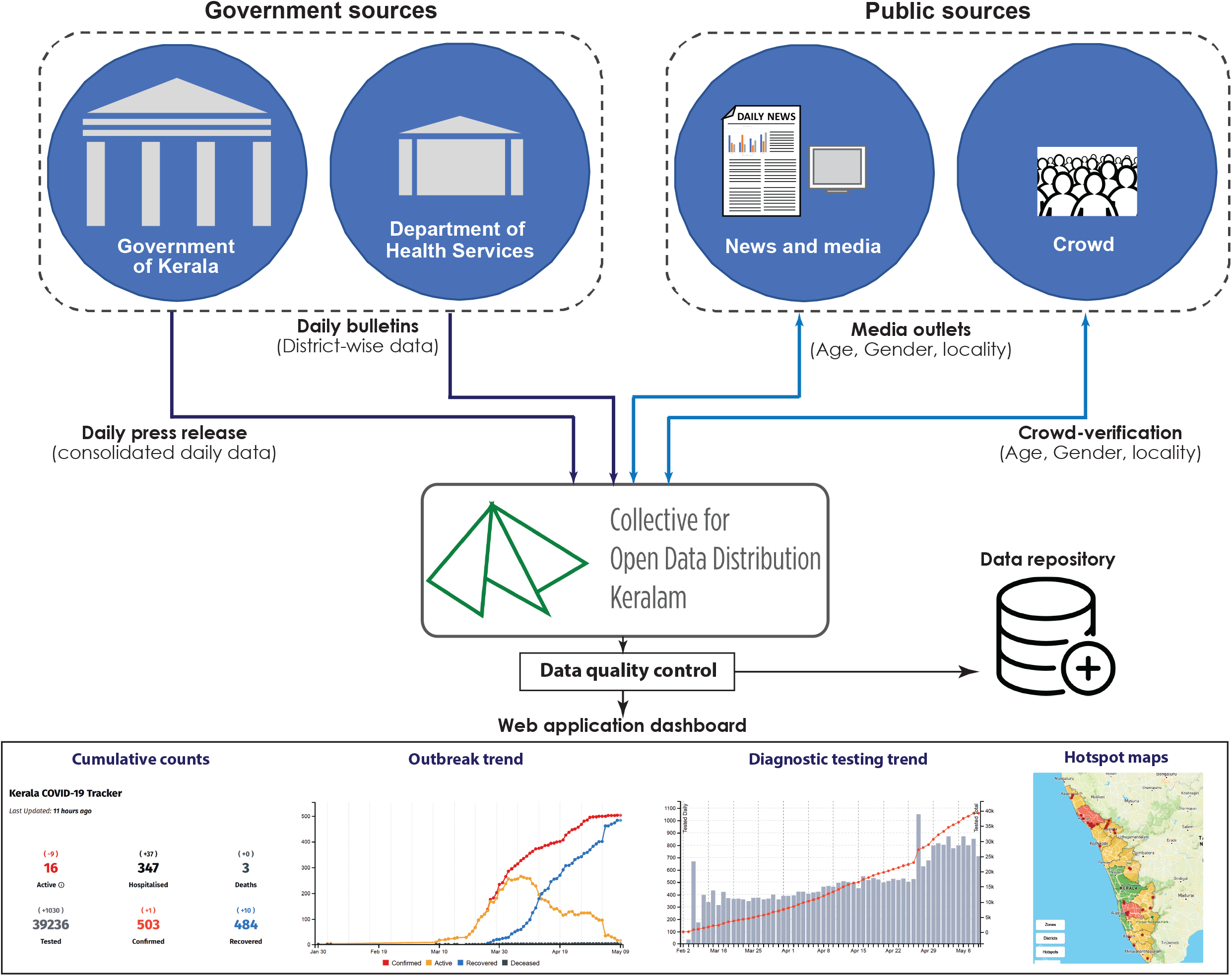
Outline of data collection, curation, and quality control for generating dataset and visualization

### Implementation of Web Application

A publicly accessible dashboard for the project is developed from a similar open-source project covid19japan.com. [29] The dashboard and related source codes are adapted and released as open-source software under MIT license, a permissive open-source software license. The dashboard has two distinctive components, a single page frontend web application accessible at https://covid19kerala.info/, and a JavaScript Object Notation (JSON) repository, which serves as an application programming interface (API) for the frontend. The API fetches data from the Google sheet and generates JSON files periodically with GitHub Actions. Both the web application and the API were created with JavaScript as the programming language and maintained using NodeJS. These portals use static-file assets without any server-side technologies. The website and the API are served through GitHub Pages, a free static web hosting service provided by GitHub (Figure 2).

**Figure 2:**
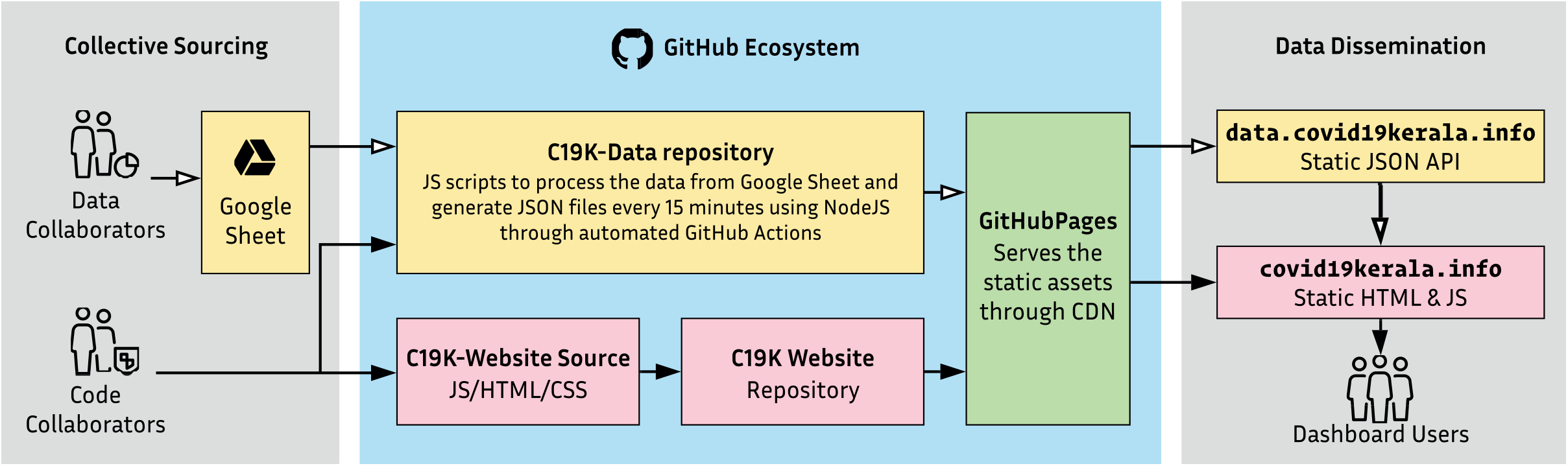
Implementation of web application and workflow

### Hotspot Mapping

COVID-19 hotspots for the Local Self Government (LSG) administration area— Panchayats, Municipalities, and Corporations were notified by the Government of Kerala, based on the recommendations (Supplementary Methods) of the Kerala State Disaster Management Authority and were updated daily through DHS bulletin as text data. A set of metadata for the LSGs, manually derived from multiple official sources with labels in both English and Malayalam, was made as an authority control for hotspots. Hotspots declared in daily bulletins are mapped to the identifiers in the LSG authority control, and containment zones were added as additional information. The LSG controlled vocabulary ensures location accuracy as well as eliminates duplicates and spelling irregularities. An independent generator periodically fetches the created hotspot list, adds spatial geometry along with the LSG metadata and generates the hotspot dataset for the dashboard. The spatial geometry of the LSGs are also manually sourced from different public resources and optimized for minimal visual indication of the boundaries of the LSGs. On the dashboard, Mapbox service renders this GeoJSON as an interactive map. [30]

## RESULTS

### Open-Data Release

The resulting open-data sets are published under Open Data Commons Attribution License v1.0 (ODC-BY 1.0). A manually curated data archive is maintained as a GitHub repository for the provenance. [31] The datasets are provided with the schema definition and an actionable data-package declaration. [32] Periodic versioned snapshots were released as ‘Covid19Kerala.info-Data’ through Zenodo (https://zenodo.org/). [33] CODD-K manages the longevity and stewardship of the data. Sufficient documentation is provided to increase the adaptability of the datasets. We ensured that the datasets complied with the Open Definition 2.1, which would enable findability, easy access, sharing, reuse, and interoperability. [26] Additionally, as per the 5-Star Linked Open Data concepts, an incremental framework for deploying data, the dataset which we sourced, enriched, and disseminated, when complied with Open Definition 2.1, evolved to 3-star open data from the 1-Star open data released by the DHS. [34] Thus, our effort by aligning to Open Definition 2.1 significantly increased openness of the data.

### Visualization of the COVID-19 data through a dashboard

Here we have collected, cleaned, and visualized publicly available data in a user-friendly bilingual progressive web application (PWA) designed to be both device and browser agnostic. For the convenience of the public, the dashboard mainly highlighted the number of individuals who are hospitalized, tested, confirmed, currently active, deceased, recovered, and people under observation (State-wise and District Data), updated daily. We also visualized maps for hotspots, and active patients, along with outbreak spread trend (new, active, and recovered cases), new cases by day, diagnostic testing trend, patients—age breakup, confirmed case trajectories at the district administration level (Figure 3A, B, and Supplementary Figure 1). To the best of our knowledge, our dashboard was the first one to be online (March 22, 2020) with a bilingual dashboard with English and the local language Malayalam, featuring outbreak map, hotspot map, and trend line map with reports of new, active, and recovered cases, along with COVID-19 related deaths in Kerala. The official dashboard version by DHS followed later. We regularly received feedback from the users and added new plots and visual tools based on user recommendations. Till June 15, 2020, the web application has seen 37,205 unique users, with an average of 2,000 visitors per day. The source code and data were open for the public to fork and analyze, thus providing a framework for a data collection, analysis, and visualization platform for future disease outbreaks.

**Figure 3:**
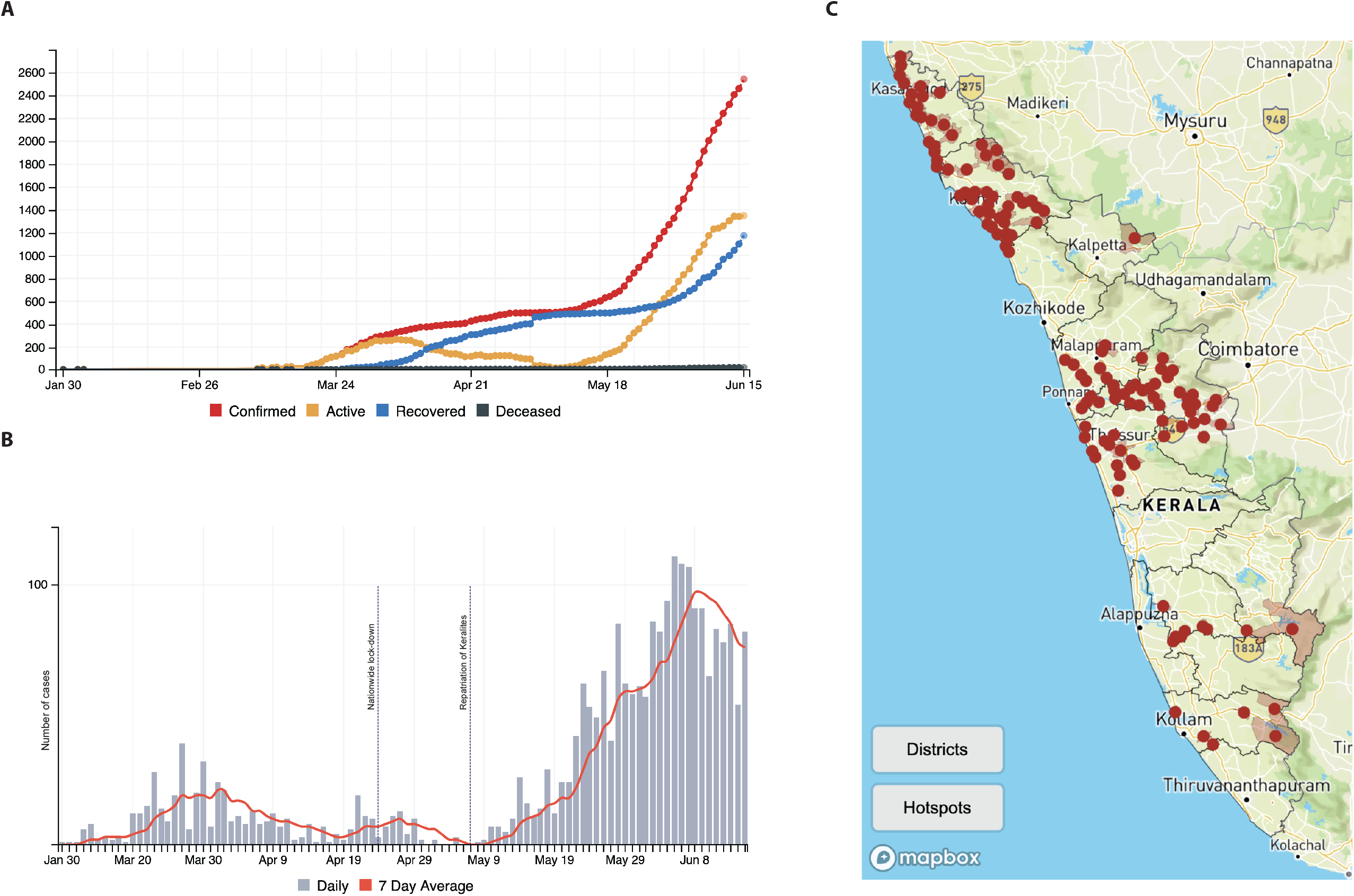
Representative images of COVID-19 outbreak trend for Kerala as visualized from the sourced data: (A) Plot showing number of confirmed, active, recovered and deceased cases (B) The trend curve, plotted with daily cases and seven days’ average is shown. The dotted lines shows the initiation of nation-wide lockdown, and repatriation of Keralites from abroad and other states (C) the hotspot map showing the districts and hotspot location

### Mapping of Hotspots for early outbreak identification

Since the SARS-CoV-2 infection outbreak occurs in clusters, early identification and isolation of these clusters are essential to contain the outbreak. Accurate tracking of the new cases and real-time surveillance is essential for the effective mitigation of COVID-19. However, the daily public bulletins by DHS did not have any unique identification code for the COVID-19 infected individuals and also for secondary contacts who have contracted the infection through contact transmission. This limited us from tracking the transmission dynamics. As an alternative, we resorted to mapping hotspots for infection as a proxy measure to indicate possible outbreak areas. Initially, red, orange, and green zones based on the number of cases were designated to each district by the Government of India. Later, the Government of Kerala started releasing COVID-19 hotspot regions of the LSG administration area. We manually curated the hotspot information from the DHS bulletins, and the dataset was published as a static JSON file in the GeoJSON format, which improves the browser caching and drops the requirement of server-sided API services. The hotspot locations were highlighted as red dots with descriptions, and when zoomed, the LSG administration area will be displayed on the map. In order to improve the visual clarity of hotspots with varying sizes of the LSGs and different zoom levels in browsers, an identifiable spot is placed on the visual center of the LSG area polygon. This inner center of the polygon was calculated with an iterative grid algorithm. To the best of our knowledge, this feature is unique to our dashboard. We also provided a toggle bar to visualize district boundaries and areas declared as hotspots at LSG resolution (Figure 3C). Owing to the lack of data, additional information such as the number of active cases in these hotspots could not be plotted.

### Outbreak trend and demographic characteristics of individuals affected with COVID-19 in Kerala from the dataset

To understand the outbreak trend and demographic characteristics of the COVID-19 infections in Kerala, we analyzed the dataset for the first 138 days of the outbreak, from January 30, 2020, to June 15, 2020. During this period, Kerala reported 2,543 cases, of which 1,174 individuals recovered during the reported period, along with 20 fatalities. Among the total number of COVID-19 infected individuals reported in Kerala, 72.36% were males, and 26.03% were females, with a large proportion of individuals falling in the age group of 20-40 (Table 1). The median age of affected individuals was 36 (0-93) (male; 38 (0-93), female; 33 (0-88)). Around 84.66% of cases had a travel history to places with reported infection, and 15.30% were infected through secondary transmission. However, even as the number of reports during this time frame increased, there was no official report of community spread. During the reported period, the state declared 163 hotspots for infection, and currently (June 15, 2020), this number has reduced to 125. Kerala has established 34 testing centers (22 government and 12 private) across the state and performed 151,686 tests during the period January 31, 2020, to June 15, 2020, which accounted for 4,359 tests per million of the population (TPR = 1.68%). In addition to routine testing, the DHS implemented additional targeted testing and testing based on random sampling in the hotspot areas. The median duration of illness was 13 days, with a trend that showed increasing recovery time for the older age group (Table 1). Oldest individuals to recover were 93 and 88 years old. The fatality rate of Kerala was 0.79%.

**Table 1:** Demographic characteristics of the individuals affected with COVID-19 in Kerala, India between January 30, 2020 to June 15, 2020

|  | All cases;<br>n (% of males) | Cases with travel history;<br>n (% of males) | Secondary transmission;<br>n (% of males) | Recovery;<br>n (% of males) | Fatality;<br>n (% of males) | Duration of illness;<br>median (range) |
| --- | --- | --- | --- | --- | --- | --- |
| Total | 2543 (72.41) | 2153 (75.6) | 389 (53.5) | 1174 (64.5) | 20 (65.0) | 13 (2-45) |
| Age break down |  |  |  |  |  |  |
| <10 | 89 (43.82) | 62 | 27 | 33 | 1 | 14 (5-32) |
| 10 - 19 | 92 (54.34) | 64 | 28 | 47 | -- | 12 (7-27) |
| 20 - 29 | 555 (69.19) | 492 | 63 | 244 | 1 | 12 (4-42) |
| 30 - 39 | 589 (78.78) | 501 | 88 | 277 | 1 | 13 (4-37) |
| 40 - 49 | 467 (79.44) | 403 | 64 | 200 | 2 | 12 (3-44) |
| 50 - 59 | 328 (76.83) | 279 | 49 | 132 | 2 | 13 (3-45) |
| 60 - 69 | 162 (76.54) | 133 | 29 | 61 | 7 | 12 (5-35) |
| 70 - 79 | 32 (50.0) | 24 | 8 | 12 | 5 | 18 (4-23) |
| > 80 | 18 (44.44) | 5 | 13 | 14 | 1 | 11 (4-41) |
| Unspecified | 211 (63.51) | 190 | 21 | 154 | -- | 15 (2-38) |
|  | All cases | Cases with travel history | Secondary transmission | Recovery | Fatality |  |
| Age;<br>median (range) | 36 (0-93) | 36 (0-83) | 36 (0-93) | 35 (2-93) | 63.5 (0-87) |  |

## DISCUSSION

In this report, we describe a citizen science initiative that leveraged publicly available unstructured COVID-19 data released daily by the Government of Kerala supplemented with news from media outlets and structured this into a knowledge bank for quick and easy interpretation through a user-friendly bilingual dashboard. The motivation for such an initiative arose due to the paucity of a real-time data visualization dashboard specific to Kerala during the initial stages of the outbreak. To the best of our knowledge, we were the first to host a visualization dashboard for COVID-19 outbreak in Kerala, with a user-friendly bilingual interface and unique features such as hotspot map. We reason that accurate information about the pandemic has made the public vigilant to adopt appropriate precautionary measures in controlling the outbreak. Our dashboard also has contributed to achieving this feat, as evidenced by the usage statistics within days of the launch. Furthermore, this open/multi-sourced dataset with a set of correlated temporal and spatial metadata was also made available for the public through an open repository, enabling retrospective analyses.

The framework developed for dataset generation and visualization can potentially be a model for advancing biomedical informatics, from a citizen-science/open data perspective. Specifically, our initiative rapidly established an easily adaptable platform and workflow for potential disease outbreaks and similar calamities, especially in resource-limited settings. With a reasonably minimal definition of data/metadata, adhering to the Open Definition 2.1, our dataset permits data-driven research on the epidemiology of the COVID-19 outbreak in Kerala and also increased openness as per 5-Star Linked Open Data concepts. Furthermore, the temporal and spatial metadata might aid in future studies involving genetic lineage diversity of SARS-CoV-2 in Kerala, in relation to the demographic characteristics and clinical phenotypes. [35 36] Thus, our model also sets an example for efficient data management in such citizen-science initiatives.

While the real-time information serves the public for assessing potential risk based on the outbreak trend/containment in a specific location; the inferences made from the emerging demographic data such as gender, age, recovery, and mortality statistics can help in refining our responses and understanding the epidemiology of COVID-19 outbreak. Also, it provides helpful insights into a rapidly developing novel pandemic for policymaking, social awareness, and enhancing compliance with the Government policies. Additionally, retrospective analysis can give insights on how policy changes or other events altered the dynamics of the COVID-19 outbreak.

Kerala has effectively utilized open/crowd-sourcing platform using citizen-led initiatives to coordinate rescue missions through social media platforms during the floods that devastated the state during 2018 and 2019. [37-39] Our collective, CODD-K evolved as a result of crowd-sourced volunteering and coordination during the floods in Kerala from 2018. Our experience during flood volunteering and the lack of appropriate data archiving during this disaster prompted us to design a real-time dashboard for COVID-19 pandemic proactively. This experience enabled us to assemble a team and launch the dashboard as rapid response during this pandemic. Experts from various domains and the general public assembled and volunteered to source data, build the dataset, visualize, distribute, and interpret the data on the outbreak through this collective. A series of recent studies involving crowd/open-source visualization of COVID-19 outbreak statistics have indicated wide popularity and impact of these community-led initiatives, including in India. [23-25] However, our approach differed from those as we sourced unstructured official data released by the government, supplemented by the information from media outlets. This strategy not only ensures authenticity but also enriches the data available in the public domain into a structured dataset, though it depends on the data release policies adopted by the different state governments. Kerala is one of the many states in India with a transparent data release policy, which ensured the authenticity of data collected through our initiative. Furthermore, the granularity of the data at the LSG levels, which are manually verified (as released in local language) gives an added advantage, in terms of data depth, over other Pan-Indian dashboards that rely on APIs to fetch cumulative data.

Although this approach seems to be efficient, an unexpected surge in cases can jeopardize the data collection, thus limiting the feasibility. During such a scenario, a trade-off between depth and breadth of data collected has to be decided. Moreover, this approach also has inherent limitations, including issues with the veracity of data, owing to the anonymity, and depth of the data released, including clinical symptoms. Since each infected case identified in Kerala was not provided with a unique ID, it was impossible to track these cases for the assessment of vital epidemiological parameters like the reproduction number (R0). Based on our experience of collating and analyzing COVID-19 data from the public domain in Kerala, we propose to frame specific guidelines for the public data release for COVID-19 or other epidemics. We recommend the release of official COVID-19 data in a consistent, structured and machine-readable format, in addition to the bulletins, which could be provided with a permanent URL and also archived in a public repository for future retrospective analyses. We also suggest releasing the assigned unique ID for the individuals affected with COVID-19, to avoid inconsistencies in reporting and to enable tracking the secondary transmission. Furthermore, providing COVID-19 associated symptomatic information, without compromising the privacy of the infected individuals will also aid in the basic understanding of the disease through analytical approaches.

Our dataset, compiled between January 30, 2020, to June 15, 2020, indicates that the infections reported in Kerala were mainly among working-age men, with a travel history of places with COVID-19 outbreak. The absence of reported community spread in this period emphasizes the effectiveness of government implemented rapid testing and quarantine measures. Active tracking and isolation of cases with travel history lead to better management with minimal COVID-19-associated death. Since the majority of cases reported in Kerala were within the age group of 20-40 years, and the patients being in constant inpatient care possibly contributed to a better outcome and lesser mortality rate, respectively. Kerala implemented vigorous COVID-19 testing, and even though the test rate was relatively low (4,359 tests per million of the population), early testing combined with strict quarantine policies for individuals with travel history prevented community spread. However, the average number of positives detected for 1,000 tests (individuals) was lesser compared to other states in India, thus negating community spread. Data from Kerala also provides insights about the mean duration of illness and the effect of increasing age on this parameter.

Collectively, we report a citizen science initiative on the COVID-19 outbreak in Kerala to collect data in a structured format utilized for visualizing the outbreak trend and describing demographic characteristics of affected individuals. While the core aim of this initiative is to document COVID-19 related information for the public, researchers, and policymakers, the implemented data visualization tool also alleviates the citizen’s anxiety around the pandemic in Kerala. We anticipate that the dataset collected will form the basis for future studies, supplemented with detailed information on clinical and epidemiological parameters from individuals with COVID-19 infection in Kerala.

## Data Availability

The dataset associated with this manuscript is made available under the Open Data Commons Attribution License v1.0 (ODC-BY 1.0)

https://zenodo.org/record/3818096

## Acknowledgments

We acknowledge Shane Reustle for his help and support for forking the Japan COVID-19 Coronavirus Tracker repository and implementation of the dashboard. We thank Jiahui Zhou for the original concept and design of the tracker. We also thank Sajjad Anwar for generously providing the administrative boundary shapefiles and geoJSONS for Kerala. Maps were generously provided by the Mapbox community team.

## Authors contribution

Conceptualization; JiU,

Data collection and curation; JiU, NN, PP, SC, SPS, MM, SJ, JeU, MK, US Formal analysis; JiU, NN, NT,

Methodology; JiU, NN, NT, SPS, AB, MK, Resources; NT, MK, AB

Software; NT, AB, MK, Supervision; JiU, STT, RE, SB

Visualization; NT, AB, PP, JiU, NN, SB Roles/Writing - original draft; SB, NNV

Writing - review & editing; SB, NNV, JiU, NN, NT

